# Social Media Reveals Psychosocial Effects of the COVID-19 Pandemic

**DOI:** 10.1101/2020.08.07.20170548

**Authors:** Koustuv Saha, John Torous, Eric D. Caine, Munmun De Choudhury

## Abstract

**Background:** The novel coronavirus disease 2019 (COVID-19) pandemic has caused several disruptions in personal and collective lives worldwide. The uncertainties surrounding the pandemic have also led to multi-faceted mental health concerns, which can be exacerbated with precautionary measures such as social distancing and self-quarantining, as well as societal impacts such as economic downturn and job loss. Despite noting this as a “mental health tsunami,” the psychological effects of the COVID-19 crisis remains unexplored at scale. Consequently, public health stakeholders are currently limited in identifying ways to provide timely and tailored support during these circumstances.

**Objective:** Our work aims to provide insights regarding people’s psychosocial concerns during the COVID-19 pandemic by leveraging social media data. We aim to study the temporal and linguistic changes in symptomatic mental health and support expressions in the pandemic context.

**Methods:** We obtain ∼60M Twitter streaming posts originating from the U.S. from 24 March-24 May 2020, and compare these with ∼40M posts from a comparable period in 2019 to attribute the effect of COVID-19 on people’s social media self-disclosure. Using these datasets, we study people’s self-disclosure on social media in terms of symptomatic mental health concerns and expressions of support. We employ transfer learning classifiers that identify the social media language indicative of mental health outcomes (anxiety, depression, stress, and suicidal ideation) and support (emotional and informational support). We then examine the changes in psychosocial expressions over time and language, comparing the 2020 and 2019 datasets.

**Results:** We find that all of the examined psychosocial expressions have significantly increased during the COVID-19 crisis – mental health symptomatic expressions have increased by ∼14%, and support expressions have increased by ∼5%, both thematically related to COVID-19. We also observe a steady decline and eventual plateauing in these expressions during the COVID-19 pandemic, which may have been due to habituation or due to supportive policy measures enacted during this period. Our language analyses highlight that people express concerns that are very specific to and contextually related to the COVID-19 crisis.

**Conclusions:** We studied the psychosocial effects of the COVID-19 crisis by using social media data from 2020, finding that people’s mental health symptomatic and support expressions significantly increased during the COVID-19 period as compared to similar data from 2019. However, this effect gradually lessened over time, suggesting that people adapted to the circumstances and their “new normal”. Our linguistic analyses revealed that people expressed mental health concerns regarding personal and professional challenges, healthcare and precautionary measures, and pandemic-related awareness. This work shows the potential to provide insights to mental healthcare and stakeholders and policymakers in planning and implementing measures to mitigate mental health risks amidst the health crisis.

## Introduction

The impacts of global public health emergencies extend beyond medical repercussions – they affect individuals and societies on many levels, causing disruptions [1,2]. In an article written by the American Psychological Association following the Ebola outbreak [3], the epidemic was described as an “epidemic of fear” – in the United States, it was described by the media as “fearbola,” to describe a paranoia that infected the country. Reports of similar feelings of anxiety, stress, and uncertainty have been repeatedly reported during other global outbreaks and pandemics [4]. The ongoing outbreak of the coronavirus, SARS-COV-2, has led to a pandemic of illness (coronavirus disease, or COVID-19) that has globally caused 16M cases and 700K deaths, reported as of the end of July 2020 [5]. According to recent surveys from the Census Bureau and the Centers for Disease Control and Prevention and other studies, the COVID-19 crisis has been reported to be associated with rapid rises in psychological distress across many nations [6], with women, the young, the less educated, and some ethnic minority groups reporting greater mental health strain [7]. On the one hand, persons are worried about the direct effects of potential infection, including fears of death, lasting disabilities, or exacerbating chronic illnesses. On the other, actions to mitigate the spread of COVID-19, including social distancing, quarantines, and business closures with resulting job losses, are a powerful source of life disruptions and emotional distress.

Fear and anxiety about a disease as infectious as COVID-19 can trigger new-onset or exacerbate existing mental illness [8]. Therefore, the practical impact of the crisis is far greater than the actual number of infection cases or fatalities [8,9]. While expressions of distress may stem from concern and worry relating to the direct impacts of the disease, they may relate as much to disruption of regular routines, sleep and eating patterns, having out-of-school children at home full-time, economic hardships and unusual volatility in financial markets, and forced geographical displacement or confinement. Indeed, some people are at risk of developing post-traumatic distress due to exposure to the multi-faceted uncertainties, or from confronting dying persons or lost loved ones. While disease mitigating efforts such as “social distancing” and “self-quarantining” are recommended [10–12] individuals in medical isolation may experience increased symptoms of anxiety and depression, as well as feelings of fear, abandonment, loneliness, and stigmatization [13,14].

Despite concerns about the myriad of social and behavioral issues associated with the COVID-19 pandemic [15,16], research has been scant to examine its psychosocial impacts or how to predict and mitigate them. Although it is anticipated that COVID-19 will have broadly ramifying effects [17,18], public health workers and crisis interventionists are limited in their ability to extend services and support in a timely, preemptive fashion. Although surveys are a step forward to support such efforts [7], due to their retrospective recall bias, limited scalability, and without being able to provide real-time insights, public health workers are not only unable to prioritize services for the most vulnerable populations, but more specifically, less equipped to direct prevention efforts towards individuals with greater propensities for adverse psychological impacts.

This paper seeks to address the above gap by drawing insights into people’s expressed mental health concerns by leveraging social media data. The rise in online and social media activity has provided an unprecedented opportunity to enhance the identification and monitoring strategies of various mental and psychosocial disorders [7,19]. Over 80% of U.S. adults use social media daily [20], placing it ahead of texting, email, and instant messaging, and disclose considerably more about themselves online than offline [21,22]. Social media provides a real-time platform where people often candidly self-disclose their concerns, opinions, and struggles during this pandemic [23]. In particular, our work targets the following research aims:

*Aim 1:* To quantitatively assess the psychosocial effects of COVID-19 pandemic using social media data.

*Aim 2:* To examine how the psychosocial effects of COVID-19 pandemic have varied over time.

*Aim 3:* To examine if social media language reflects the major psychosocial concerns during the COVID-19 pandemic.

For the above research questions, we measure psychosocial effects in terms of symptomatic mental health expressions of anxiety, depression, stress, and suicidal ideation, and expressions seeking emotional and informational support. Our work is founded on a large body of work on studying mental health and psychosocial dynamics with social media data [24–30]. Several studies have leveraged Twitter (which is also the data of our current study) to study health attributes and public health [30], including symptoms related to diseases [31], disease contagion [32], obesity and physical health [33], mood and depressive disorders [28,34], mental health self-disclosures [27], post-traumatic stress disorder [35], addictive behaviors and substance use [36,37], etc. Because social media data (and Twitter posts in particular) are recorded in-the-moment they provide rich information about both the individual as well as the larger world [30]. In particular, we draw on two kinds of prior work --- on symptomatic mental health expressions and on support expressions. Related to the former, Saha et al. (2019), in their study on the effects of psychiatric medications on Twitter, developed classifiers of mental health symptomatic expressions using social media language, which we replicate in our current work [25]. Related to the latter, we draw upon Sharma and De Choudhury (2018) and Saha and Sharma (2020) developed classifiers of social support expressions, specifically emotional and informational support [38,39].

## Methods

### Data

To study people’s psychosocial expressions on social media, we obtain Twitter data. Twitter is one of the most popular social media platforms, and its public-facing, micro-blogging based design enables candid self-disclosure and self-expressions for individuals [27].

### Twitter Streaming API

We collect data in our study using the Twitter Streaming API. Twitter Streaming API is an official data collection API that Twitter shares with researchers providing free access to 1% sample of its data on a set of parameters set by researchers. That is, for a given set of parameters, Twitter queries the volume of available data at a particular moment [40]. If the volume of the query exceeds 1% of all Twitter posts at that moment, then the response is sampled to be less than 1%. However, the Twitter Streaming API is like a black-box with a lack of transparency in the sampling methodology [41,42], yet, this is one of the few forms of unfettered and large-scale social media data access to researchers outside social media companies [43], and has also used in a number of prior work, including health [35,44,45]. The Discussion section revisits the limitations of our study due to the challenges of Twitter Streaming API.

For the purposes of our study, we use two kinds of parameters, 1) language of a Twitter post as “english”, and 2) geo-location bounds set to be within the geographic coordinates of U.S. Therefore, our ensuing analyses concern Twitter data that at least fulfill both these criteria. We note that the location filter additionally prevents any retweets in the dataset as retweets are not geo-location labeled by design on Twitter [40], allowing us to study only originally created Twitter posts.

### *Treatment* Data

In particular, we focus our study on the U.S. population and leverage the Twitter Streaming API. Using geo-bounded coordinates, we collect 1% of real-time Twitter data originating from the U.S. We collect 59,096,694 Twitter posts between March 24, 2020, and May 24, 2020. Because this dataset comes from the same period when the COVID-19 outbreak occurred, we label this dataset as the *Treatment* dataset. We note that this period saw an exponential growth in reported COVID-19 infection cases (∼50K to ∼1M) and fatalities (∼1K to ∼56K) in the U.S. [46]. During these two months, federal and state policies and laws were enacted to control or mitigate the spread of the outbreak, including school and work closures, stay-at-home orders, and Coronavirus Aid, Relief, and Economic Security Act [47].

### *Control* Data

To understand the social media expressions particularly attributed to the COVID-19 crisis, we obtain a control dataset that originates from the same geographical location (U.S.) and similar time period, but from the previous year (2019). Prior work [47,48] motivates this approach of obtaining control data that acts as a baseline and likely minimizes confounding effects due to geo-temporal seasonality in lifestyle, activities, experiences, and unrelated events that may have some psychosocial bearing. We obtain a similarly-sized dataset of 40,875,185 Twitter posts shared between March 24, 2019, and May 24, 2019.

Both the above Treatment and Control datasets have been collected in real-time and therefore, they are the entire 1% sample of Twitter posts returned in real-time, and we do not conduct any additional sampling on this data. We note that the size of Control data is smaller than that of Treatment [42]despite each consisting of the same two months duration. This could be because the volume of Twitter posts in [40,42]the entire Twitter increased significantly in 2020 [49] leading to an increase in 1% sample as well. However, we cannot make any such conclusion, especially because of the lack of transparency in how Twitter conducts the 1% sampling [42].

## Psychosocial Effects of COVID-19

Towards our first research aim of understanding the psychosocial impacts of the COVID-19 outbreak, we conduct two types of analysis on our Twitter dataset, which we describe below. Our work builds upon the vast, rapidly growing literature studying mental health concerns and psychosocial expressions within social media data [19,21,24–28,34,48,50–52].

### Symptomatic Mental Health Expressions

Drawing on the work referenced above, we hypothesize that people’s self-disclosure expressions on social media can reveal symptomatic mental health expressions attributed to the COVID-19 crisis. We examine symptomatic expressions of anxiety, depression, stress, and suicidal ideation. These are not only some of the most critical mental health concerns but also have been attributed to be consequences of the pandemic outbreak [15,53,54].

To identify mental health symptomatic expressions in social media language, Saha et al. (2019) built machine learning classifiers [25] using transfer learning methodologies --- the main idea here is to infer mental health attributes in an unlabeled data by transferring a classifier trained on a different labeled dataset. These classifiers are *n*-gram (*n*=1,2,3) based binary Support Vector Machine (SVM) models where the positive class of the training datasets stems from appropriate Reddit communities (*r/depression* for depression, *r/anxiety* for anxiety, *r/stress* for stress, and *r/SuicideWatch* for suicidal ideation), and the negative class of training datasets comes from non-mental health-related content on Reddit — a collated sample of 20M posts, gathered from 20 subreddits from the landing page of Reddit during appropriately the same period as the mental health subreddit posts, such as *r/AskReddit, r/aww, r/movies*, and others. These classifiers perform at a high accuracy of approximately 0.90 on average on held-out test data [25].

#### Clinical Validity

Saha et al.’s classifiers, used here, have also been shown to transfer well on Twitter with an 87% agreement between machine-predicted labels and expert appraisal [25,48],, where experts annotated posts in the classification test data using DSM-5 [55] criteria of mental health symptoms. Bagroy et al. [55,56] reported additional validation of such derived insights with feedback from clinical experts. In this work, the outcomes of the mental health expression classifiers were compared with those given by human coders on the same (random) sample of social media posts; the latter coded the posts based on a codebook developed using prior qualitative and quantitative studies of mental health disclosures on social media and literature in psychology on markers of mental health expressions. Coders not only agreed with the outcomes of the classifiers (Cohen’s κ was 0.83), but also noted that the classifiers could identify explicit expressions of first-hand experience of psychological distress or mental health concerns (“i get overwhelmingly depressed”) as well as expressions of support, help, or advice around difficult life challenges and experiences (“are there any resources I can use to talk to someone about depression?”). Further details about these classifiers, including their detailed performance, predictive features demonstrating model interpretability, and efficacy of transfer to Twitter data, may be found in [25,48,56]. We use these classifiers to machine label both our *Treatment* and *Control* datasets. Table 1 shows example Twitter posts in our dataset which exhibit mental health symptomatic expressions (because many of these labels are comorbid, we show example posts that exhibit one or more of these mental health symptomatic expressions).

**Table 1.**
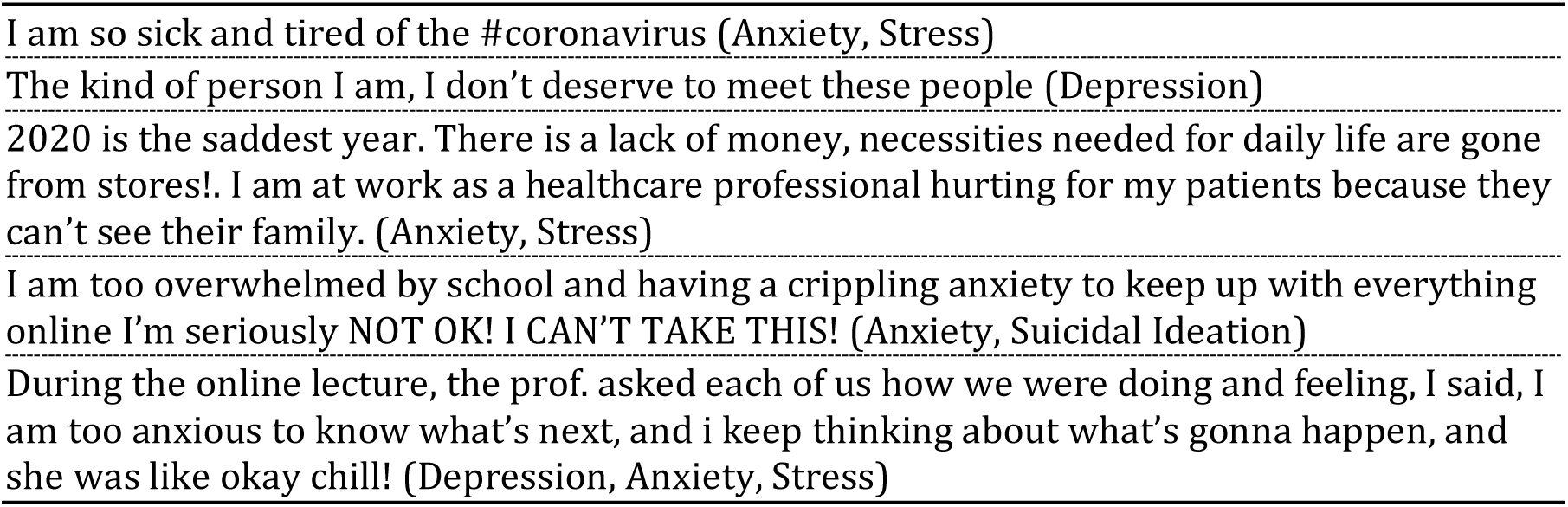
Example paraphrased posts in the Treatment data that exhibit high symptomatic mental health expressions.

|  |
| --- |
| I am so sick and tired of the #coronavirus (Anxiety, Stress) |
| The kind of person I am, I don't deserve to meet these people (Depression) |
| 2020 is the saddest year. There is a lack of money, necessities needed for daily life are gone from stores!. I am at work as a healthcare professional hurting for my patients because they can't see their family. (Anxiety, Stress) |
| I am too overwhelmed by school and having a crippling anxiety to keep up with everything online I'm seriously NOT OK! I CAN'T TAKE THIS! (Anxiety, Suicidal Ideation) |
| During the online lecture, the prof. asked each of us how we were doing and feeling, I said, I am too anxious to know what's next, and i keep thinking about what's gonna happen, and she was like okay chill! (Depression, Anxiety, Stress) |

### Support Expressions

Social support is considered an essential component in helping people cope with psychological distress [57]. Research reports that supportive interactions can even have a “buffering effect” [57,58]; that is, they can be protective against the negative consequences of mental health. With the wide adoption of web and social media technologies, support-seeking (and providing) is increasingly happening online and has been shown to be efficacious [21,59]. In fact, a meta-analysis indicates that online support is effective in decreasing depression and increasing self-efficacy and quality of life [60]. In the context of suicide, certain types of social support in Reddit communities may reduce the chances of future suicidal ideation among those seeking mental health help [61]. Oh et al. further showed that surveyed Facebook users demonstrate a positive relationship between having health concerns and seeking health-related social support [62]. Indeed, during global crises such as COVID-19, when many of the physical sites for healthcare (including mental health) have been closed or have very restricted access, it is likely that online support has proliferated [63]. Fear of potential infection may further have alienated individuals in need to pursue formal treatment, therapy, and support, perhaps channelizing their support seeking efforts online and on social media.

According to the “Social Support Behavioral Code” [64], two forms of support that have received theoretical and empirical attention are emotional and informational support. Emotional support (ES) corresponds to empathy, encouragement, and kindness, while informational support (IS) corresponds to information, guidance, and suggestions [38,65]. These two forms of support have been found to be most prevalent and effective in several studies of online support and social media [38,62,66,67]. Social media enables individuals to self-disclose and express in making their emotional and informational needs known and sought [67]. Andalibi et al. found that these two kinds of support can co-occur with other forms of support, such as posts seeking emotional support often seek esteem and network support [66], and Attai et al. noted that Twitter is effective in seeking and providing health-related informational needs [68], contextually related with our problem of interest.

To identify support expressions on social media, we use an expert-appraised dataset and classifier built in prior work [38,39]. These are binary SVM classifiers identifying the degree (high/low) of ES and IS in social media posts. When the predictions of these classifiers were cross-validated with expert annotations from Sharma and De Choudhury’s data [38], the classifiers were found to have *k*-fold cross-validation accuracies of 0.71 and 0.77 in ES and IS classifications respectively [39]. Similar to the symptomatic expressions classifiers, the classifiers of support expressions are transferred from Reddit and typically perform well in our dataset due to the high linguistic equivalence between Reddit and Twitter datasets [34]. We further manually inspect a random set of 125 Twitter posts in our dataset using the methods outlined in prior work [25,56] to rate each Twitter post with binary high or low ES and IS. We find that the manual ratings and classifier ratings show a high agreement of 88% and 93%, respectively, indicating statistical significant transfer classification on Twitter. We use these classifiers to label the presence of ES and IS in our *Treatment* and *Control* datasets. Table 2 shows a few example paraphrased posts of support expressions in our *Treatment* dataset.

**Table 2.**
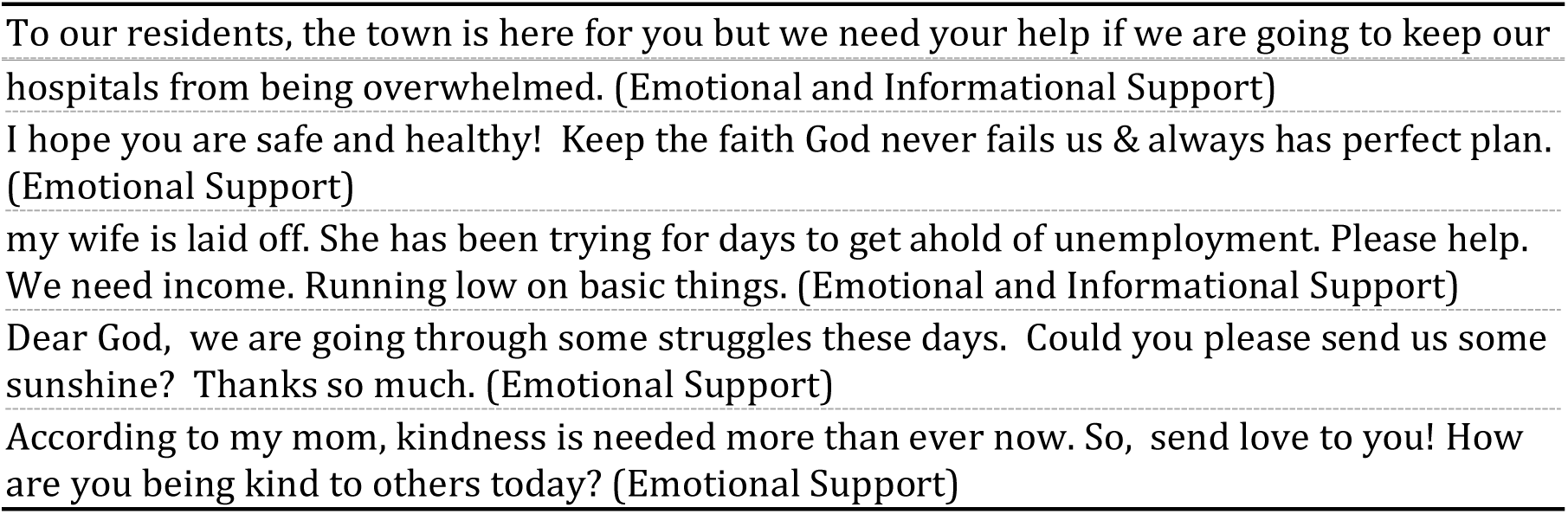
Example paraphrased posts in the *Treatment* data on support expressions

## Examining Psychosocial Expressions over Time and Language

Next, we describe methods to examine how the COVID-19 pandemic may have caused changes in psychological expressions by comparing our *Treatment* (outbreak year) and *Control* (no-outbreak year) datasets. For both our datasets, we aggregate the number of posts that express symptomatic and support expressions by day and by type. We compare the pervasiveness of each kind of measure in the datasets, along with conducting statistical significance in their differences using 2-sample t-tests and effect sizes (Cohen’s d).

### Temporal Variation

For our second research aim, we compare the daily variation of measures between *Treatment* and *Control* datasets, we transform our data into standardized *z*-scores. Our datasets rely on the Twitter streaming API, and are subject to daily inconsistencies of available data each day [41]. Transformed *z*-scores are not sensitive to such absolute values and inconsistencies, and essentially quantify the number of standard deviations by which the value of the raw score is above or below the mean. Similar standardization techniques have been adopted in prior social media time-series studies [48,69]. *z*-scores are calculated as *(x−μ)/σ*, where *x* is the raw value, *μ* is the mean and σ is the standard deviation of the population. Here, to obtain population *μ* and *σ*, in addition to our *Treatment* and *Control* data, we also include a year-long Twitter data of over 240M Twitter posts (September 2018 to August 2019). For each of the measures in symptomatic and support expressions, we first calculate *μ* and *σ* on the per-day occurrence of that particular measure in the dataset of over 300M Twitter posts (combining 240M posts between September 2018 and August 2019 and 60M posts in Treatment data between March and May 2020). Then, for each measure, we calculate the *z*-score per day and interpret positive *z*-scores as values above the mean, and negative *z*-scores as those below the mean.

### Linguistic Differences

For our third research aim, we examine COVID-19 related linguistic differences in the psychosocial expressions on social media, we employ an unsupervised language modeling technique, the Sparse Additive Generative Model (SAGE) [70]. Given any two datasets, SAGE selects salient keywords by comparing the parameters of two logistically parameterized-multinomial models using a self-tuned regularization parameter to control the tradeoff between frequent and rare keywords. We conduct SAGE to identify distinguishing n-grams (*n*=1,2,3) between the *Treatment* and *Control* datasets, where each n-gram is returned with a SAGE score. The magnitude of an *n*-gram’s SAGE score signals the degree of its “uniqueness” or saliency, and in our case a positive SAGE score (above 0) indicates that the n-gram is more salient in the *Treatment* data, whereas a negative SAGE score (below 0) denotes greater saliency in the *Control* data.

SAGE allows us to obtain how the expressions differ during the COVID-19 outbreak as compared to the *Control* period. We conduct two SAGE analyses, one each for symptomatic and support expressions. For the symptomatic expressions, we first obtain posts that are indicative of either of anxiety, depression, stress, or suicidal ideation in *Treatment* and *Control*, and obtain SAGE for the two datasets. We do similar for support expressions by obtaining posts that are indicative of either emotional or informational support.

Finally, we cross-examine the salient keywords across symptomatic and support expressions, to study how concerns are prevalent in either or both of the kinds of expressions. We measure log-likelihood ratios (*LLR*) along with add-1 smoothing, where *LLR* close to 0 indicates comparable frequencies, *LLR*<1 indicates the greater frequency in symptomatic expressions and *LLR*>1 indicates the greater frequency in support expressions. Together these linguistic analyses enable us to obtain psychological concerns, and understand how COVID-19 has psychosocially affected individuals, and to contextualize these concerns in the literature on consequences of global crises.

## Results

We summarize our first set of results in Table 1. For all our measures, we find statistical significance (as per *t*-tests) as well as significant effect sizes (Cohen’s d > for all measures [71]) in social media expressions in the *Treatment* data as compared to that in *Control*. Assuming that most other confounders were minimized due to the geo-temporal similarity of the datasets, our findings indicate that the COVID-19 outbreak led to an increase in people’s symptomatic and support expressions of mental health. We elaborate on the results below.

## Temporal Variation

Figure 1a shows the changes in symptomatic mental health expressions for the same period in *Treatment* (2020) and *Control* (2019) years. We find that the *Treatment* and *Control* show significant differences in the people’s symptomatic expressions (Table 3), among which, anxiety shows the most significant increase (21.32%), followed by suicidal ideation (19.73%), depression (10.18%), and stress (3.76%). Figure 1b shows the evolution of support expressions change in the *Treatment* and *Control* datasets. Like above, the differences are significant (Table 3), and we find that not only emotional support increases by 4.77%, and informational support also increases by 4.78%.

**Table 3.** Comparing social media expressions in *Treatment* (2020) and *Control* (2019) (*p<0.05, **p<0.01, ***p<0.001).

|  | **Treatment (2020)** |  | **Control (2019)** |  |  |  |  |
| --- | --- | --- | --- | --- | --- | --- | --- |
| **Expression** | **Mean** | **Stdev.** | **Mean** | **Stdev.** | **$\Delta\%$** | **Cohen's d** | **t-stat.** |
| *Symptomatic Mental Health Expressions* |  |  |  |  |  |  |  |
| Anxiety | 1.65 | 0.20 | 1.35 | 0.08 | 21.32 | 1.96 | 12.60*** |
| Depression | 9.00 | 0.60 | 8.17 | 0.35 | 10.18 | 1.71 | 10.72*** |
| Stress | 19.31 | 0.77 | 18.61 | 0.43 | 3.76 | 0.81 | 3.65** |
| Suicidal Ideation | 3.14 | 0.31 | 2.62 | 0.13 | 19.73 | 2.14 | 13.54*** |
| *Support Expressions* |  |  |  |  |  |  |  |
| Emotional Support | 8.56 | 0.84 | 8.17 | 0.50 | 4.77 | 0.46 | 2.87** |
| Informational Support | 1.75 | 0.18 | 1.67 | 0.08 | 4.78 | 0.56 | 3.58*** |

**Figure 1.**
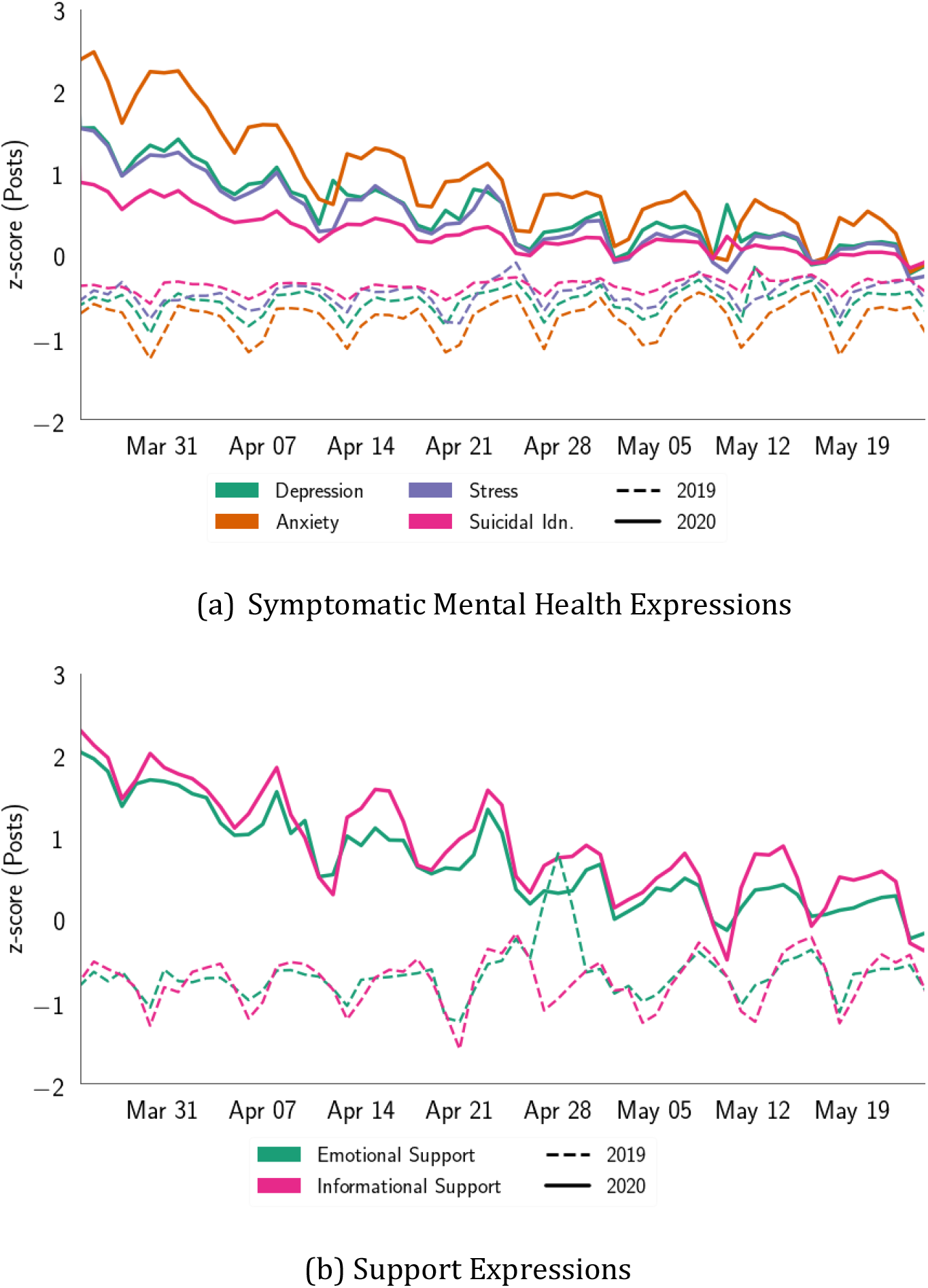
Comparison of symptomatic mental health and support expressions on social media posts in the same period (March 25 - May 25) in 2019 and 2020 (COVID-19 outbreak year).

In both the plots of Figure 1, we find a general trend of negative slope (avg. slope = - 0.03) within the *Treatment* year, which is closer to zero slope (avg. slope = 3.19*10−4) in the *Control* dataset. This may suggest that within the *Treatment* year, people’s mental health expressions gradually leveled out over time, despite the growing rate of COVID-19 active cases. The plots indicate that psychological expressions almost converge at the tails. This could likely be due to people’s habituation with the situation and surroundings with the passage of time [70,72], as has been observed for other crisis events [48,73]; however, this needs to be explored further. Within the *Control* dataset, we observe a sudden peak on April 28, 2019, which could be attributed to a shooting incident at a synagogue in San Diego [74]. The observations reflect that the COVID-19 pandemic has increased people’s mental health expressions on social media, aligning with other contemporary literature and media reports [8,53].

## Linguistic Expressions

### Symptomatic Mental Health Expressions

Table 4 summarizes the language differences as per SAGE for posts expressing high mental health expressions in *Treatment* and *Control* periods --- keywords with positive SAGE saliently occur in *Treatment*, whereas those with negative SAGE saliently occur in the *Control* data. A majority of the keywords that occur in the *Treatment* period are contextually related to the COVID-19 pandemic, such as *covid19, coronavirus, social distancing, stayathome isolation*. These keywords are used in posts expressing mental health concerns either explicitly, e.g., “Social distancing is both sad and anxiety-inducing at the same moment”, or implicitly, e.g., “In order to get my family treated, I will do more than beg, and I will donate 25K for research to develop COVID19 vaccine.” We also find that the *Treatment* period uses keywords referring to key personnel such as *dr fauci* (referring to Anthony Fauci, one of the leads in the incumbent White House Coronavirus Task Force in the U.S. and Director of the National Institute of Allergy and Infectious Diseases since 1984 [75]) and political figures like *Nury Martinez* and *Donald Trump*. Further, we find keywords, such as *essential workers, doctor jobs*, and *risking lives*, which describe high-risk worker situations, e.g., “I am not complaining about going to work, rather, I am concerned about risking my health for work.”, and certain treatment suggestions that evolved during this period [76] such as *garlic, malaria*, and *hydroxychloroquine*, e.g., “I do worry tho!He is focused on job numbers, approval? ratings and repeating mistruths. His spouting of 2 drugs, one for malaria & the other a Z-pack.A Senior couple tried these untested drugs; wife is in ICU & husband also hospitalized! This is irresponsible & dangerous!”

**Table 4.**
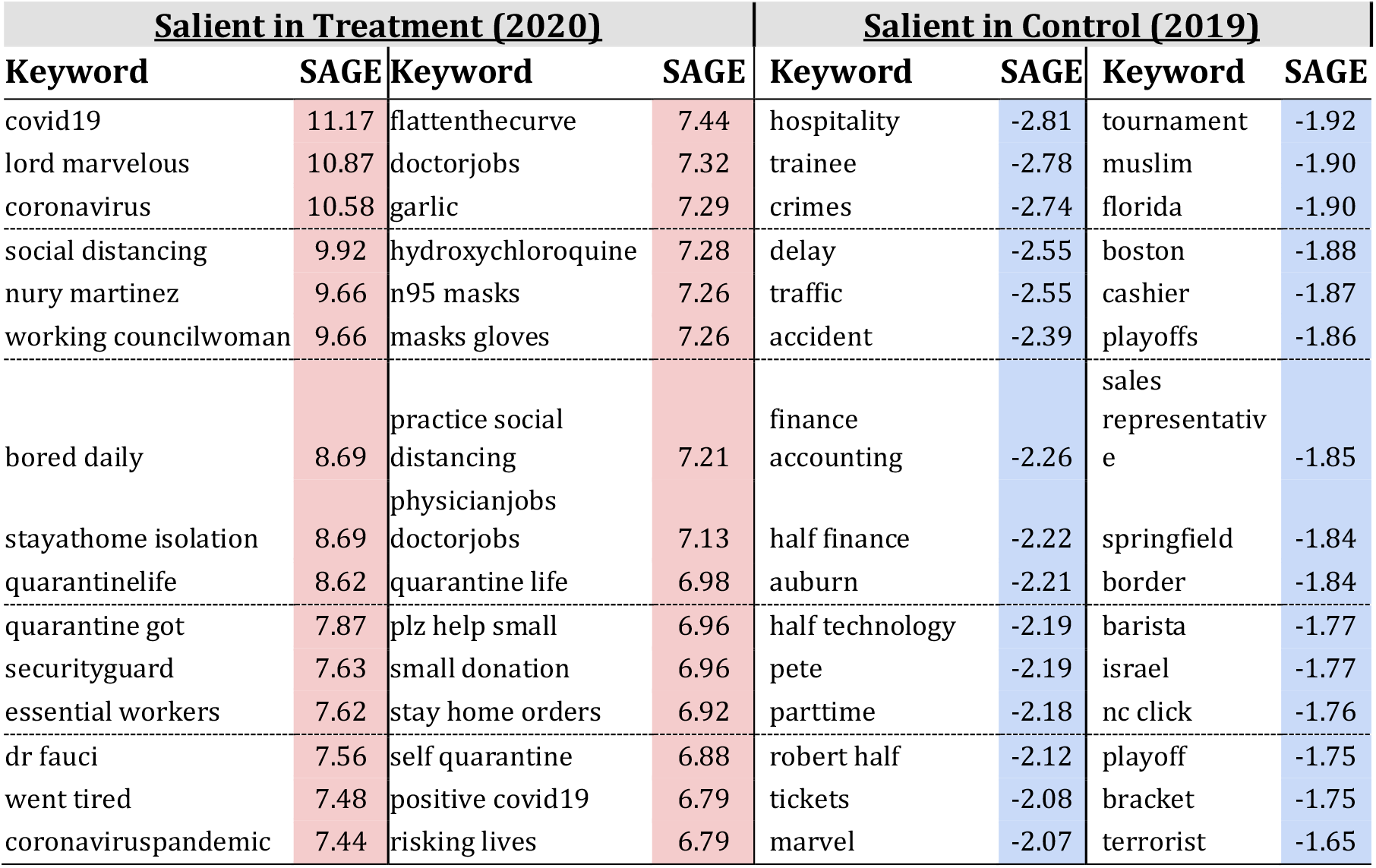
Top salient n-grams (n=1.2,3) for symptomatic mental health concerns in *Treatment* and *Control* datasets (SAGE Analysis [70]). Positive/ red-shaded SAGE scores indicate greater saliency in Treatment (2020) data, and Negative/ blue-shaded SAGE scores indicate greater saliency in Control (2019) data.

### Support Expressions

Table 5 lists the top keywords as per SAGE for support posts in *Treatment* and *Control* period. Like above, keywords with positive SAGE saliently occur in *Treatment*, whereas those with negative SAGE saliently occur in the *Control* data; we find keywords that explicitly relate to COVID-19 occur in the *Treatment* period. We also find that the *Treatment* period consists of posts that seek support related to job and pay such as, *losing jobs, need pay*, and *furloughed*, e.g., “Many in our community have lost their jobs, are underinsured and are struggling to make ends meet. Providing pantries, hot meals, hotspots and distance learning opportunities is now more critical than ever, please donate.” Our data also reveals the prevalence of contextually related keywords such as *masks, ppe, hoarding, stockpile*, and *sanitizer* that are medically recommended prevention and containment measures of COVID-19 infection, e.g., “Please contact me if you have any N95 mask or know to obtain some. My sister and a few friend work in the OR and they do not have the supplies to stay safe, they have patients who have #COVID19. TY! #HealthcareHeroes.”

**Table 5.** Top salient n-grams (n=1.2,3) for support expressions in Treatment and *Control* datasets (SAGE Analysis [70]). Positive/ red-shaded SAGE scores indicate greater saliency in Treatment (2020) data, and Negative/ blue-shaded SAGE scores indicate greater saliency in Control (2019) data.

| Salient in Treatment (2020) |  |  |  | Salient in Control (2019) |  |  |  |
| --- | --- | --- | --- | --- | --- | --- | --- |
| Keyword | SAGE | Keyword | SAGE | Keyword | SAGE | Keyword | SAGE |
| lord | 7.93 | staysafe | 4.67 | hospitality | -2.86 | cashier | -1.79 |
| fauci | 6.70 | food bills | 4.66 | duke | -2.51 | springfield | -1.79 |
| ventilators | 6.59 | disinfectant | 4.64 | shift supervisor | -2.24 | delay | -1.76 |
| quarantine | 6.47 | hand sanitizer | 3.08 | tampa | -2.21 | barista store | -1.76 |
| securityofficer | 6.11 | clorox | 3.03 | advisor | -1.95 | boston | -1.76 |
| n95 | 5.53 | medical supplies | 2.97 | customerservice | -1.92 | counter | -1.75 |
| hope staying safe | 5.36 | trying times | 2.89 | investigation | -1.89 | barista | -1.74 |
| ppe | 5.25 | risking lives | 2.87 | manager retail | -1.87 | columbia | -1.73 |
| wearing masks | 5.20 | stockpile | 2.86 | traffic | -1.87 | meeting retail | -1.73 |
| uncertain times | 5.16 | father passed | 2.36 | muslim | -1.86 | informational meeting | -1.73 |
| healthcare workers | 5.01 | hoarding | 2.31 | store manager | -1.85 | stlouis | -1.72 |
| furloughed | 5.00 | mask | 2.31 | tickets | -1.85 | marvel | -1.70 |
| asymptomatic | 4.95 | medical professionals | 2.27 | playoffs | -1.83 | marketing | -1.68 |
| people quarantine | 4.90 | losing jobs | 2.27 | cubs | -1.82 | server | -1.67 |
| fighting stigma | 4.82 | toilet paper | 2.05 | border | -1.81 | accident | -1.64 |

### Linguistic Comparability

Finally, Table 6 shows the results of the lexical comparability analysis, where log likelihood ratios (*LLRs*) demarcate the top keywords used for symptomatic mental health expressions and support expressions within the *Treatment* dataset. We find that keywords, such as safety precautions (*wear masks*), healthcare and treatment (*health care workers, hospitalized, beds*, and *icu*), and life/death (*passed away, kill people, human lives*, and *deaths*) comparably overlap in both kinds of psychological expressions (*LLR∼*0). These keywords are also used to raise awareness and express solidarity with healthcare and high-risk workers, e.g., “Taking all safety precautions and adhering to the guidelines established by our health care professionals will keep us safe.” Our lexico-psychological analyses reveal that more clinically relevant keywords and symptoms occur frequently in symptomatic expressions (*LLR*>0), e.g., *sleep schedule* and *tested positive*, whereas, socially relevant and stressful circumstances are more prevalent in support expressions (*LLR*<0), e.g., *im single parent, starve*, and *lost jobs*.

**Table 6.** Distribution of social media keywords across high symptomatic mental health and support expressions within *Treatment* period using Log Likelihood Ratios (*LLR*). Keywords with *LLR*>0 distinctly occur in high symptomatic expressions, those with <0 distinctly occur in support expressions, and those ∼0 occur comparably in both.

| <u>**LLR>0**</u> |  | <u>**LLR~0**</u> |  | <u>**LLR<0**</u> |  |
| --- | --- | --- | --- | --- | --- |
| **Keyword** | **LLR** | **Keyword** | **LLR** | **Keyword** | **LLR** |
| sleep schedule | 0.75 | infected | -0.01 | im single parent | -1 |
| lonely | 0.64 | wear masks | -0.01 | starve | -1 |
| anxiety | 0.62 | need help | -0.01 | meditate | -1 |
| isolation | 0.56 | kill people | -0.01 | sorry loss | -0.73 |
| stay safe | 0.56 | need pay | 0 | care people | -0.7 |
| bored | 0.56 | health care workers | 0 | hard times | -0.45 |
| tested positive | 0.52 | passed away | 0 | people sick | -0.4 |
| quarantine life | 0.52 | seriousness | 0 | helping people | -0.4 |
| homeschooling | 0.51 | human lives | 0 | sorry hear | -0.39 |
| tired | 0.5 | deaths | 0 | urged | -0.33 |
| doctor | 0.48 | domestic violence | 0 | new yorkers | -0.29 |
| fighting stigma | 0.46 | comforting | 0 | lost jobs | -0.21 |
| depression | 0.45 | hospitalized | 0 | hope family | -0.21 |
| stuck inside | 0.42 | beds | 0 | selfish | -0.21 |
| sane | 0.41 | icu | 0.01 | desperate | -0.21 |

## Discussion

### Principal Results

Our study suggests that social media posts during the COVID-19 pandemic contain a significantly higher frequency of symptomatic mental health and support expressions than a comparable dataset from the same period in the previous year. The effect sizes and statistical differences observed in our analyses provide evidence that the COVID-19 may have led to mental health concerns compared to other normative times. We also find that they topically relate to the ongoing crisis situation, and include concerns such as: treatment, precautionary measures, loss of jobs, school closings, stockpiling of basic livelihood necessities, feeling lonely, bored, and tired of the restrictions and constraints put on by the ongoing pandemic, and so on. Our findings suggest that although the COVID-19 pandemic has amplified mental health risks and concerns, it may have heightened a sense of belonging and solidarity among individuals — bringing them together, raising collective awareness, and encouraging them to provide support to one another. We also find expressions of solidarity with healthcare and high-risk workers, suggesting that people have been considerate about these workers, who have expressed desire to set up opportunities for donating to those who have lost jobs during the crisis --- this also aligns with recent media reports and WHO guidelines of tackling the pandemic [77,78]. Media reports have also indicated how benevolent neighbors have been tending to their elderly neighbors by delivering their groceries and other basic necessities [79].

However, mental health experts say that while the crisis is amplifying risk factors for suicide, the coronavirus outbreak’s effect on individuals’ mental and emotional wellbeing is complex [80]. Suicide is multifaceted, and while economic loss is a risk factor, so are depression, isolation and fear of the future. At the same time, the crisis is possibly creating a sense of belonging for individuals at risk for suicide as stress and anxiety are normalized, and people come together to better support one another during a crisis [81,82]. Our data shows a significant impact of COVID-19 on suicidal ideation which calls for enhanced importance about population-scale mental healthcare, such as using approaches like universal screening (i.e., Zero Suicide Model)[83]. As Florida noted in a recent article [84]: “The long-term toll on mental health of social isolation, remote work, and economic insecurity could have impacts akin to post-traumatic stress disorder; yet, the new focus on mental health may reduce stigma and increase the availability of support services.” Indeed, the world beyond the crisis may be one in which mental health is more honestly recognized and supported.

Interestingly, we note that our findings indicate a gradual leveling out of these expressions — both symptomatic and supportive, may reflect a developing ‘new normal.’ In February 2020, it seemed unthinkable the white-collar workforce of many countries would soon be working solely from home, it seemed unthinkable air travel would plummet by 96%, and all major sporting events will be called off. Indeed, epidemiologists surmise that many if not most of the changes surrounding the rhythms of our daily life are likely to fade over time, just as they did after the 1918 influenza epidemic [84]. In other words, the pandemic would make us revisit and possibly reform many of our lifestyle choices and civic roles, and the persistent discussion of the ‘new normal’ may help bring order to our current turbulence. Others have argued that perhaps the crisis is an prelude to a ‘new paradigm,’ as recently noted by the World Economic Forum [84,85]: “Feeling unsettled, destabilized and alone can help us empathize with individuals who have faced systematic exclusions long-ignored by society even before the rise of COVID-19 – thus stimulating urgent action to improve their condition.” We should therefore “revel in the discomfort of the current moment to generate a ‘new paradigm,’ not a ‘new normal.’ The leveling out trend in our data gives empirical ground to these conjectures.

Nevertheless, if robust antiviral treatments are developed and rolled out relatively quickly and/or if a vaccine becomes available soon enough, presumably, the changes will be short-lived, and the new normal may be temporary. But if the pandemic comes back in larger waves over the next few seasons, like was the case with historical epidemics, the economic, political, and social crises that have arisen as a consequence will lead to deeper ramifications in turn leading to longer-lasting or permanent changes. Future research will need to explore the persistence of the new normal and the emergence of a possible new paradigm as the pandemic evolves, and therein the mental health impacts further along in the crisis. A study like ours on the ongoing pandemic is a step towards leveraging large-scale online data to understand people’s response to the crisis, and thinking about means to address the major concerns. Our work bears implications in digital technology driven mental health interventions to provide tailored support to people’s concerns during the crisis --- as a recent work by Rudd and Beidas pointed out four point guidelines to build innovative and expansive solutions towards improving public mental health [86]. The variety of concerns and help-seeking factors reflected in our study can also help several stakeholders ranging across mental health facilitators and policy makers towards early preparedness and interventions for mental health support. Similarly, our methods can offer the potential to build public health surveillance technologies that surface early warning signs of the effects of the various events related to the pandemic and other crises. The potential of social media to assist in the response to the pandemic is clear but also dependent on the accuracy of underlying methods. The reach of social media allows for broad access that transcends national borders or cultural differences. Using this access to meet the increasing need for help seeking --- online and social media data is in a prime position to offer people personalized guidance towards accurate information, health care resources, and even basic lifestyle interventions. Underlying this potential is thus the ability of social media data to classify the state and needs of each individual, and use that information to tailor a customized response. Precedents for such a system are abound as seen in several prior work [27–31,37,87–89].

## Comparison with Prior Work

COVID-19 is not the first pandemic — catastrophic pandemics have been occurring at regular intervals throughout human history, with the 1918 influenza epidemic being the last one before the current pandemic [84,85,90]. The backdrop of the 1918 pandemic was that it happened just before the advent of modern psychiatry as a science and a clinical specialty – a time when psychoanalysis was gaining recognition as an established treatment within the medical community [91,92]. Consequently, psychiatry has had little opportunity to consider such historically important phenomena through its clinical, scientific lens, until now. Although outbreaks of the Zika and Ebola virus, MERS, and SARS managed to draw global attention, stirring up anxiety and uncertainty in societies, scholars have noted that participation of mental health experts in pandemic preparedness has remained negligible [93]. Consequently, our ability to understand mental health responses as well as the mental health burden in pandemic outbreaks have been limited [94]. For instance, a routinely practiced method of infection control, quarantine and social distancing have received surprisingly little attention in psychiatric literature so far. Baumeister and Leary (1995) [95] contended that humans need frequent contacts, and crisis events further stimulate a need for affiliation and intimacy. Therefore, prolonged isolation and separation from families and their community can have profound effects on individuals even if they are not directly affected by the disease [4]. In the current pandemic, the additional layer of extensive use of social media and exposure to often sensationalized online news, while in physical isolation, may add new complexities to implementing emotional epidemiology in managing concerns, fears, and misconceptions [96,97], as these tools have been argued to bear negative effects on psychological wellbeing [97],[98].

By adopting social media as a lens to unpack these previously less understood dimensions of a pandemic’s mental health effects, our work is one step towards closing some of the above-noted gaps. The published literature posits that the distress and anxiety among individuals in this COVID-19 pandemic may increase the incidence of mental disorders [53,54,99]; data thus far from the U.S. point to a population increase in psychological distress of 10% compared to 2018 data [8], a trend which is in line with our present results. These rates may be higher in those regions heavily exposed to COVID-19 or among individuals working during the pandemic, with a recent review reporting over 20% prevalence of anxiety, also consistent with our findings [8].

Prior work found that mental health discourse on Twitter ranges across stigmatizing, inspirational, resource, medical, and social dimensions of expressions [100], and our study revealed similar topical diversity in our dataset. Further, we detected through social media many of the stresses associated with the pandemic – e.g., prolonged isolation, exposure to pandemic-related death, loss of income/career, increased workload, and lack of pertinent and accurate information. These results align with epidemiological findings that COVID-19 has led elevated mental health symptoms for individuals: Nelson et al. (2020) surveyed two thousand individuals from U.S., Canada, and Europe and found elevated symptoms of anxiety and depression compared to historical norms, and observed factors similar to the concerns we detected regarding symptomatic expressions and those related to seeking support. They also reinforce the summary data released by the Crisis Text Line (a major crisis helpline in the U.S.) listing major concerns of crisis support sought during this period [101] — 80% conversations mentioning *“virus”*, 34% mentioning *“anxiety”*, 34% feeling solidarity with friends and family, etc. Along similar lines, there have been numerous reports about the increasing number of mental health crisis helpline calls during this period [102,103], providing further support and external validation that our social media findings reflect many of the same elements of distress expressed offline during this crisis.

Next, our temporal analyses pointed to a steady decline in people’s expressed psychosocial concerns during the two month study period (Figure 1), which conforms with similar findings in Google search queries as stay-at-home orders and other COVID-19 related policy changes were implemented in the U.S. [104]. We note contemporary social computing research studying various aspects of the social media discourse related to COVID-19 [63,105–107]. By providing complementary evidence to observations by Mackey et al. [105] and Stokes et al. [106] on expressed (mental health) concerns during the crisis, our work further underscores their findings using a comparable (control) dataset, reinforcing and providing empirical credibility to the impression that the COVID-19 pandemic has indeed caused or contributed directly to the mental health concerns that we describe.

## Limitations and Future Work

We note some limitations in our work, many of which present excellent directions for future research. We recognize the lack of transparency related to the Twitter streaming API. Recent research has also questioned the credibility of the “1% Twitter stream” aspect noting that actual sampling data is smaller than what it ideally should have been [41]. Given these data limitations, we decided against conducting several descriptive and fine-grained analyses (such as comparing regions), and refrained from making claims based on comparing absolute numbers of those impacted by various mental health concerns. For example, we cannot define based on our data, whether there were increased or decreased Twitter postings during our COVID-19 study period compared to the same months in 2019. Again, we chose to filter English-only Twitter posts given both algorithmic limitations of our methodologies, and lower prevalence of non-English data (particularly in the U.S. context). However, future work can extend our methodologies to conduct analyses in other languages to draw richer insights.

Despite the strengths of Twitter as a data source that provides us unobtrusive access to large-scale, unstructured, and naturalistic data of people’s candid self-disclosure, and that it has been a valuable source to study disaster and crisis response [108], we acknowledge that this data inherently suffers from many biases, such as self-selection and representation [109]. We can only study those who self-select to express on Twitter. Pew surveys report that social media platforms are under-representative of minorities, although Twitter is an exception, which over-represents minorities such as Blacks, Hispanics, and women [20]. There is already a digital divide in terms of social media use where the population is skewed towards young adults and white-collar workers. Further, technology and social media could be a luxury to marginalized and underprivileged populations, and any sort of technology-driven support and assistance will disproportionately affect different individuals based on technology use [88]. Similarly, a single platform cannot provide a complete picture; different platforms (such as Facebook, Reddit, Twitter, instant messaging services) have unique design strengths and weaknesses both in terms of their affordances, as well as who uses them. Therefore, as highlighted in a recent article by Chunara and Cook (2020), public health surveillance (including that for COVID-19) can account for several factors such as the “population at risk” in epidemiology and demographic disparities in the use and behavioral expressions on social media [23].

We understand that our work is observational, and as any other observational study, does not measure “true causality”. Watts (2014) noted the impossibility to test all explanations and confounders simultaneously [110]. However, by including and comparing against a control data, we minimize geo-temporal and seasonal confounds, thereby enabling us to provide stronger evidence and insights than purely correlational analyses regarding the effects of COVID-19 pandemic on people’s mental health. We also note that support expressions in our study can not only include support-seeking but also support-providing expressions. This has also enabled us to observe how solidarity and sense of belonging proliferated during the COVID-19 crisis. Future work can build separate high precision classifiers for each kind of expression to disentangle the prevalence of seeking and providing expressions during the crisis.

Further, while we did have data beyond May 24, 2020, we decided to exclude those in order to keep our focus on the effects on social media expressions due to COVID-19 and minimize those that followed the death of George Floyd on May 25, 2020, in the light of the Black Lives Matter protests throughout the U.S. [111]. We also are aware that, with the continuing nature of the pandemic, our conclusions are restricted to the mental health and support-seeking concerns expressed during a finite study period. Events since the end of the study period underscore the dynamic nature of these events, as different parts of the U.S. are heavily affected, while others are recovering and some remain relatively spared. It will be important to extend this work temporally, increase the size of future samples, and, whenever possible, add geospatial specificity to future analyses. The latter will be especially important for potential supportive interventions locally, if one has the resources and the ability to assemble recurring, near-real-time local “snapshots” as a basis for community focused preventive interventions. Further, our analyses can be extended to retrospectively examine the aftereffects of particular global and local events (such as policy changes) related to the pandemic.

## Conclusion

Our work, like those of others studying other major events, further reinforces the potential utility of accessing and analyzing social media data in near-real-time to ‘take the temperature’ of communities. This will require a more focused and robust collection of locally targeted information to build samples that are sufficiently large to produce reliably representative datasets to be useful for public health interventions. Further work is now needed to track mental health-related expressions and those reflecting needs for support throughout the pandemic, which has seen dynamic changes associated with disease spread to areas that had been less affected during the early months of the outbreak. This geospecific research may further enhance our understanding of the causal connections between COVID-spread and waves of expressed distressed. Having the ability to present locally pertinent, contemporaneous analyses offers the opportunity for local public health and mental health providers, as well as political leaders, to develop and deploy targeted support services in a timely fashion.

## Data Availability

The data is public social media data. We will make the Twitter IDs available.

## Acknowledgments

MDC was partly supported by a COVID-19 related Rapid Response Research (RAPID) grant #2027689 from the National Science Foundation.

## Abbreviations

COVID-19: Coronavirus Disease 2019
API: Application Programming Interface

## References

1. Hays JN. Epidemics and Pandemics: Their Impacts on Human History. ABC-CLIO; 2005. ISBN:9781851096589

2. Boscarino JA. Community Disasters, Psychological Trauma, and Crisis Intervention. Int J Emerg Ment Health 2015;17(1):369–371. PMID:25983663

3. Lu S. An epidemic of fear [Internet]. PsycEXTRA Dataset. 2015. [doi:10.1037/e520422015-003]

4. Morganstein JC, Fullerton CS, Ursano RJ, Donato D, Holloway HC. Pandemics: Health Care Emergencies [Internet]. Textbook of Disaster Psychiatry. p. 270–284. [doi: 10.1017/9781316481424.019]

5. COVID-19 Coronavirus Pandemic [Internet]. Worldometers. [cited 2020 Jul 25]. Available from: https://www.worldometers.info/coronavirus/

6. Basu T. The coronavirus pandemic is a game changer for mental health care [Internet]. Technology Review. [cited 2020 Jul 25]. Available from: https://www.technologyreview.com/2020/03/20/905184/coronavirus-online-therapy-mental-health-app-teletherapy/

7. Resnick B. A third of Americans report anxiety or depression symptoms during the pandemic [Internet]. Vox. [cited 2020 Jul 25]. Available from: https://www.vox.com/science-and-health/2020/5/29/21274495/pandemic-cdc-mental-health

8. McGinty EE, Presskreischer R, Han H, Barry CL. Psychological Distress and Loneliness Reported by US Adults in 2018 and April 2020. JAMA [Internet] 2020 Jun 3; PMID:32492088

9. Holmes EA, O’Connor RC, Hugh Perry V, Tracey I, Wessely S, Arseneault L,Ballard C, Christensen H, Silver RC, Everall I, Ford T, John A, Kabir T, King K, Madan I, Michie S, Przybylski AK, Shafran R, Sweeney A, Worthman CM, Yardley L, Cowan K, Cope C, Hotopf M, Bullmore E. Multidisciplinary research priorities for the COVID-19 pandemic: a call for action for mental health science [Internet].The Lancet Psychiatry. 2020. p. 547–560. [doi: 10.1016/s2215-0366(20)30168-1]

10. Organization WH, World Health Organization. Operational considerations for case management of COVID-19 in health facility and community. Interim guidance [Internet]. Pediatria i Medycyna Rodzinna. 2020. p. 27–32. [doi: 10.15557/pimr.2020.0004]

11. Hellewell J, Abbott S, Gimma A, Bosse NI, Jarvis CI, Russell TW, Munday JD, Kucharski AJ, Edmunds WJ, Centre for the Mathematical Modelling of Infectious Diseases COVID-19 Working Group, Funk S, Eggo RM. Feasibility of controlling COVID-19 outbreaks by isolation of cases and contacts. Lancet Glob Health 2020 Apr;8(4):e488–e496. PMID:32119825

12. Saxena SK. Coronavirus Disease 2019 (COVID-19): Epidemiology, Pathogenesis, Diagnosis, and Therapeutics. Springer Nature; 2020. ISBN:9789811548147

13. Miller G. Social distancing prevents infections, but it can have unintended consequences [Internet]. Science. 2020. [doi: 10.1126/science.abb7506]

14. Varatharaj A, Thomas N, Ellul MA, Davies NWS, Pollak TA, Tenorio EL, Sultan M, Easton A, Breen G, Zandi M, Coles JP, Manji H, Al-Shahi Salman R, Menon DK, Nicholson TR, Benjamin LA, Carson A, Smith C, Turner MR, Solomon T, Kneen R, Pett SL, Galea I, Thomas RH, Michael BD, CoroNerve Study Group. Neurological and neuropsychiatric complications of COVID-19 in 153 patients: a UK-wide surveillance study. Lancet Psychiatry [Internet] 2020 Jun 25; PMID:32593341

15. Pfefferbaum B, North CS. Mental Health and the Covid-19 Pandemic. N Engl J Med 2020 Aug 6;383(6):510–512. PMID:32283003

16. Fiorillo A, Gorwood P. The consequences of the COVID-19 pandemic on mental health and implications for clinical practice. Eur Psychiatry 2020 Apr 1;63(1):e32. PMID:32234102

17. Heymann DL, Shindo N. COVID-19: what is next for public health? [Internet]. The Lancet. 2020. p. 542–545. [doi: 10.1016/s0140-6736(20)30374-3]

18. Weinstock CP. Ripple effects of covid-19 strain mental health systems [Internet]. US News. [cited 2020 Jul 25]. Available from: http:usnews.com/news/healthiest-communities/articles/2020-06-04/coronavirus-ripple-effects-strain-mental-health-systems

19. Chancellor S, De Choudhury M. Methods in predictive techniques for mental health status on social media: a critical review. NPJ Digit Med 2020 Mar 24;3:43. PMID:32219184

20. Social Media Fact Sheet [Internet]. PEW. [cited 2020 Jul 25]. Available from: http:pewinternet.org/fact-sheet/social-media

21. Andalibi N, Haimson OL, De Choudhury M, Forte A. Understanding Social Media Disclosures of Sexual Abuse Through the Lenses of Support Seeking and Anonymity [Internet]. Proceedings of the 2016 CHI Conference on Human Factors in Computing Systems. 2016. [doi: 10.1145/2858036.2858096]

22. Qiu L, Lin H, Leung AK, Tov W. Putting their best foot forward: emotional disclosure on Facebook. Cyberpsychol Behav Soc Netw 2012 Oct;15(10):569– 572. PMID:22924675

23. Chunara R, Cook SH. Using Digital Data to Protect and Promote the Most Vulnerable in the Fight Against COVID-19. Front Public Health 2020 Jun 12;8:296. PMID:32596201

24. Guntuku SC, Schwartz HA, Kashyap A, Gaulton JS, Stokes DC, Asch DA, Ungar LH, Merchant RM. Variability in Language used on Social Media prior to Hospital Visits. Sci Rep 2020 Mar 12;10(1):4346. PMID:32165648

25. Saha K, Sugar B, Torous J, Abrahao B, Kiciman E, De Choudhury M. A Social Media Study on the Effects of Psychiatric Medication Use. Proc Int AAAI Conf Weblogs Soc Media 2019 Jun 7;13:440–451. PMID:32280562

26. Saha K, Kim SC, Reddy MD, Carter AJ, Sharma E, Haimson OL, De Choudhury M. The Language of LGBTQ Minority Stress Experiences on Social Media [Internet]. Proceedings of the ACM on Human-Computer Interaction. 2019. p. 1–22. [doi: 10.1145/3361108]

27. Ernala SK, Labetoulle T, Bane F, Birnbaum ML, Rizvi AF, Kane JM, De Choudhury M. Characterizing audience engagement and assessing its impact on social media disclosures of mental illnesses. In Twelfth International AAAI Conference on Web and Social Media 2018, June.

28. De Choudhury M, Gamon M, Counts S, Horvitz E. Predicting Depression via Social Media. ICWSM 2013.

29. Saha K, Weber I, Birnbaum ML, De Choudhury M. Characterizing Awareness of Schizophrenia Among Facebook Users by Leveraging Facebook Advertisement Estimates. J Med Internet Res 2017 May 8;19(5):e156. PMID:28483739

30. Dredze M. How Social Media Will Change Public Health [Internet]. IEEE Intelligent Systems. 2012. p. 81–84. [doi: 10.1109/mis.2012.76]

31. Paul MJ, Dredze M. You are what you Tweet: Analyzing Twitter for public health. International AAAI Conference on Weblogs and Social Media 2011.

32. Sadilek A, Kautz H, Silenzio V. Predicting disease transmission from geo-tagged micro-blog data. Twenty-Sixth AAAI Conference on Artificial Intelligence 2012.

33. Gore RJ, Diallo S, Padilla J. You Are What You Tweet: Connecting the Geographic Variation in America’s Obesity Rate to Twitter Content. PLoS One 2015 Sep 2;10(9):e0133505. PMID:26332588

34. Saha K, Chan L, De Barbaro K, Abowd GD, De Choudhury M. Inferring Mood Instability on Social Media by Leveraging Ecological Momentary Assessments [Internet]. Proceedings of the ACM on Interactive, Mobile, Wearable and Ubiquitous Technologies. 2017. p. 1–27. [doi: 10.1145/3130960]

35. Coppersmith G, Harman C, Dredze M. Measuring Post Traumatic Stress Disorder in Twitter. Eighth international AAAI conference on weblogs and social media 2014.

36. Kiciman E, Counts S, Gasser M. Using longitudinal social media analysis to understand the effects of early college alcohol use. ICWSM 2018.

37. Liu J, Weitzman ER, Chunara R. Assessing Behavioral Stages From Social Media Data. CSCW Conf Comput Support Coop Work 2017 Feb;2017:1320–1333. PMID:29034371

38. Sharma E, De Choudhury M. Mental Health Support and its Relationship to Linguistic Accommodation in Online Communities [Internet]. Proceedings of the 2018 CHI Conference on Human Factors in Computing Systems - CHI’18. 2018. [doi: 10.1145/3173574.3174215]

39. Saha K, Sharma A. Causal Factors of Effective Psychosocial Outcomes in Online Mental Health Communities. In Proceedings of the International AAAI Conference on Web and Social Media 2020, May;14:590–601.

40. Filter realtime Tweets [Internet]. Developer Twitter API. [cited 2020 Aug 17]. Available from: https://developer.twitter.com/en/docs/twitter-api/v1/tweets/filter-realtime/guides/basic-stream-parameters

41. Pfeffer J, Mayer K, Morstatter F. Tampering with Twitter’s Sample API[Internet]. EPJ Data Science.2018. [doi: 10.1140/epjds/s13688-018-0178-0]

42. Morstatter F, Pfeffer J, Liu H. When is it biased? [Internet]. Proceedings of the 23rd International Conference on World Wide Web - WWW’14 Companion. 2014. [doi: 10.1145/2567948.2576952]

43. Boyd D, Crawford K. CRITICAL QUESTIONS FOR BIG DATA [Internet].Information, Communication & Society. 2012. p. 662–679. [doi:10.1080/1369118x.2012.678878]

44. Burton SH, Tanner KW, Giraud-Carrier CG, West JH, Barnes MD. “Right time, right place” health communication on Twitter: value and accuracy of location information. J Med Internet Res 2012 Nov 15;14(6):e156. PMID:23154246

45. Tsuya A, Sugawara Y, Tanaka A, Narimatsu H. Do cancer patients tweet? Examining the twitter use of cancer patients in Japan. J Med Internet Res 2014 May 27;16(5):e137. PMID:24867458

46. Centers for Disease Control and Prevention [Internet]. CDC. [cited 2020 May 9]. Available from: cdc.gov

47. Timeline of the COVID-19 pandemic in the United States [Internet]. Wikipedia. [cited 2020 Jul 25]. Available from: http:en.wikipedia.org/wiki/Timeline_of_the_COVID-19_pandemic_in_the_United_States

48. Saha K, De Choudhury M. Modeling Stress with Social Media Around Incidents of Gun Violence on College Campuses [Internet]. Proceedings of the ACM on Human-Computer Interaction. 2017. p. 1–27. [doi: 10.1145/3134727]

49. Iqbal M. Twitter Revenue and Usage Statistics (2020) [Internet]. Business of Apps. 2020 [cited 2020 Aug 17]. Available from: https://www.businessofapps.com/data/twitter-statistics/#2

50. De Choudhury M, Kiciman E, Dredze M, Coppersmith G, Kumar M. Discovering Shifts to Suicidal Ideation from Mental Health Content in Social Media. Proc SIGCHI Conf Hum Factor Comput Syst 2016 May;2016:2098–2110. PMID:29082385

51. Guntuku SC, Russell Ramsay J, Merchant RM, Ungar LH. Language of ADHD in Adults on Social Media [Internet]. Journal of Attention Disorders. 2019. p. 1475–1485. [doi: 10.1177/1087054717738083]

52. Saha K, Weber I, De Choudhury M. A Social Media Based Examination of the Effects of Counseling Recommendations After Student Deaths on College Campuses. Proc Int AAAI Conf Weblogs Soc Media 2018 Jun;2018:320–329. PMID:30505628

53. Bedford J, Enria D, Giesecke J, Heymann DL, Ihekweazu C, Kobinger G, Clifford Lane H, Memish Z, Oh M-D, Sall AA, Schuchat A, Ungchusak K, Wieler LH. COVID-19: towards controlling of a pandemic [Internet]. The Lancet. 2020. p. 1015–1018. [doi: 10.1016/s0140-6736(20)30673-5]

54. Taha S, Matheson K, Cronin T, Anisman H. Intolerance of uncertainty, appraisals, coping, and anxiety: The case of the 2009 H1N1 pandemic [Internet]. British Journal of Health Psychology. 2014. p. 592–605. [doi: 10.1111/bjhp.12058]

55. American Psychiatric Association. Diagnostic and Statistical Manual of Mental Disorders (DSM-5®). American Psychiatric Pub;2013. ISBN:9780890425572

56. Bagroy S, Kumaraguru P, De Choudhury M. A Social Media Based Index of Mental Well-Being in College Campuses. Proc SIGCHI Conf Hum Factor Comput Syst 2017 May;2017:1634–1646. PMID:28840202

57. Kummervold PE, Gammon D, Bergvik S, Johnsen J-AK, Hasvold T, Rosenvinge JH. Social support in a wired world: Use of online mental health forums in Norway [Internet]. Nordic Journal of Psychiatry. 2002. p. 59–65. [doi: 10.1080/08039480252803945]

58. Cohen S, Wills TA. Stress, social support, and the buffering hypothesis [Internet]. Psychological Bulletin. 1985. p. 310–357. [doi: 10.1037/0033-2909.98.2.310]

59. De Choudhury M, De S. Mental health discourse on reddit: Self-disclosure, social support, and anonymity. In Eighth international AAAI conference on weblogs and social media 2014, May.

60. Rains SA, Young V. A Meta-Analysis of Research on Formal Computer-Mediated Support Groups: Examining Group Characteristics and Health Outcomes [Internet]. Human Communication Research. 2009. p. 309–336. [doi: 10.1111/j.1468-2958.2009.01353.x]

61. De Choudhury M, Kiciman E. The Language of Social Support in Social Media and its Effect on Suicidal Ideation Risk. Proc Int AAAI Conf Weblogs Soc Media 2017 May;2017:32–41. PMID:28840079

62. Oh HJ, Lauckner C, Boehmer J, Fewins-Bliss R, Li K. Facebooking for health: An examination into the solicitation and effects of health-related social support on social networking sites [Internet]. Computers in Human Behavior. 2013. p. 2072–2080. [doi: 10.1016/j.chb.2013.04.017]

63. Luo C, Li Y, Chen A, Tang Y. What triggers online help-seeking retransmission during the COVID-19 period? Empirical evidence from Chinese social media [Internet]. [doi: 10.1101/2020.06.13.20130054]

64. Cutrona CE, Suhr JA. Controllability of Stressful Events and Satisfaction With Spouse Support Behaviors [Internet]. Communication Research. 1992. p. 154– 174. [doi: 10.1177/009365092019002002]

65. Nambisan P. Information seeking and social support in online health communities: impact on patients’ perceived empathy. J Am Med Inform Assoc2011 May 1;18(3):298–304. PMID:21486888

66. Andalibi N, Haimson OL, De Choudhury M, Forte A. Social Support, Reciprocity, and Anonymity in Responses to Sexual Abuse Disclosures on Social Media [Internet]. ACM Transactions on Computer-Human Interaction. 2018. p. 1–35. [doi: 10.1145/3234942]

67. Wang Y-C, Kraut R, Levine JM. To stay or leave? [Internet]. Proceedings of the ACM 2012 conference on Computer Supported Cooperative Work - CSCW’12. 2012. [doi: 10.1145/2145204.2145329]

68. Attai DJ, Cowher MS, Al-Hamadani M, Schoger JM, Staley AC, Landercasper J. Twitter Social Media is an Effective Tool for Breast Cancer Patient Education and Support: Patient-Reported Outcomes by Survey. J Med Internet Res 2015 Jul 30;17(7):e188. PMID:26228234

69. Golder SA, Macy MW. Diurnal and seasonal mood vary with work, sleep, and daylength across diverse cultures. Science 2011 Sep 30;333(6051):1878–1881. PMID:21960633

70. Eisenstein J, Ahmed A, Xing EP. Sparse additive generative models of text.In Proceedings of the 28th International Conference on International Conference on Machine Learning 2011, June.

71. Cohen J. Statistical Power Analysis for the Behavioral Sciences [Internet]. 2013. [doi: 10.4324/9780203771587]

72. Rankin CH, Abrams T, Barry RJ, Bhatnagar S, Clayton DF, Colombo J, Coppola G, Geyer MA, Glanzman DL, Marsland S, McSweeney FK, Wilson DA, Wu C-F, Thompson RF. Habituation revisited: an updated and revised description of the behavioral characteristics of habituation. Neurobiol Learn Mem 2009 Sep;92(2):135–138. PMID:18854219

73. Choudhury MD, De Choudhury M, Monroy-Hernández A, Mark G. “Narco” emotions [Internet]. Proceedings of the 32nd annual ACM conference on Human factors in computing systems - CHI’14. 2014. [doi: 10.1145/2556288.2557197]

74. Medina J, Mele C, Murphy H. One Dead in Synagogue Shooting Near San Diego; Officials Call It Hate Crime. New York Times 2019 Apr;

75. Anthony Fauci, MD [Internet]. NIH. Available from: https://niaid.nih.gov/about/anthony-s-fauci-md-bio

76. Colson P, Rolain J-M, Lagier J-C, Brouqui P, Raoult D. Chloroquine and hydroxychloroquine as available weapons to fight COVID-19. Int J Antimicrob Agents. 2020. p. 105932. PMID:32145363

77. Mental health and psychosocial considerations during the COVID-19 outbreak [Internet]. World Health Organization. [cited 2020 Jul 20]. Available from: https://apps.who.int/iris/bitstream/handle/10665/331490/WHO-2019-nCoV-MentalHealth-2020.1-eng.pdf

78. Broom D. A pandemic of solidarity? This is how people are supporting one another as coronavirus spreads [Internet]. World Economic Forum. 2020 [cited 2020 Aug 18]. Available from: https://www.weforum.org/agenda/2020/03/covid-19-coronavirus-solidarity-help-pandemic/

79. Free C. People across the country are delivering groceries free. It’s a solidarity, not charity [Internet]. Washington Post. [cited 2020 Jul 25]. Available from: https://www.washingtonpost.com/lifestyle/2020/04/27/people-across-country-are-delivering-groceries-free-its-solidarity-not-charity/

80. Reger MA, Stanley IH, Joiner TE. Suicide Mortality and Coronavirus Disease 2019—A Perfect Storm? [Internet]. JAMA Psychiatry. 2020. [doi: 10.1001/jamapsychiatry.2020.1060]

81. Cohn MA, Mehl MR, Pennebaker JW. Linguistic markers of psychological change surrounding September 11, 2001. Psychol Sci 2004 Oct;15(10):687–693. PMID:15447640

82. González-Sanguino C, Ausín B, Castellanos MÁ, Saiz J, López-Gómez A, Ugidos C, Muñoz M. Mental health consequences during the initial stage of the 2020 Coronavirus pandemic (COVID-19) in Spain. Brain Behav Immun 2020 Jul;87:172–176. PMID:32405150

83. Labouliere CD, Vasan P, Kramer A, Brown G, Green K, Kammer J, Finnerty M, Stanley B. «Zero Suicide» – A model for reducing suicide in United States behavioral healthcare [Internet]. Suicidologi. 2018. [doi: 10.5617/suicidologi.6198]

84. Florida R. Beyond covid-19 lies a new normal– and new opportunities [Internet]. Bloomberg. [cited 2020 Jul 25]. Available from: https://www.bloomberg.com/news/features/2020-06-25/the-new-normal-after-the-coronavirus-pandemic

85. Asonye C. There’s nothing new about the ‘new normal’. Here’s why [Internet]. Weforum. [cited 2020 Jul 25]. Available from: https://www.weforum.org/agenda/2020/06/theres-nothing-new-about-this-new-normal-heres-why/

86. Rudd BN, Beidas RS. Digital Mental Health: The Answer to the Global Mental Health Crisis? JMIR Ment Health 2020 Jun 2;7(6):e18472. PMID:32484445

87. Yoo DW, Birnbaum ML, Van Meter AR, Ali AF, Arenare E, Abowd GD, De Choudhury M. Designing a Clinician-Facing Tool for Using Insights From Patients’ Social Media Activity: Iterative Co-Design Approach [Internet]. JMIR Mental Health. 2020. p. e16969. [doi: 10.2196/16969]

88. Pendse SR, Lalani FM, De Choudhury M, Sharma A, Kumar N. “Like Shock Absorbers”: Understanding the Human Infrastructures of Technology-Mediated Mental Health Support [Internet]. Proceedings of the 2020 CHI Conference on Human Factors in Computing Systems. 2020. [doi: 10.1145/3313831.3376465]

89. Saha K, Bayraktaroglu AE, Campbell AT, Chawla NV, De Choudhury M, D’Mello SK. Social media as a passive sensor in longitudinal studies of human behavior and wellbeing. Extended Abstracts of the 2019 CHI Conference on Human Factors in Computing Systems 2019.

90. Cunha BA. Influenza: historical aspects of epidemics and pandemics [Internet]. Infectious Disease Clinics of North America. 2004. p. 141–155. [doi: 10.1016/s0891-5520(03)00095-3]

91. Aassve A, Alfani G, Gandolfi F, Le Moglie M. Epidemics and trust: the case of the spanish flu. 2020;

92. Phillips H. The Recent Wave of “Spanish” Flu Historiography [Internet]. Social History of Medicine. 2014. p. 789–808. [doi: 10.1093/shm/hku066]

93. Huremović D. Psychiatry of Pandemics: A Mental Health Response to Infection Outbreak. Springer; 2019. ISBN:9783030153465

94. Ornell F, Schuch JB, Sordi AO, Kessler FHP. “Pandemic fear” and COVID-19: mental health burden and strategies. Braz J Psychiatry 2020 Apr 3;42(3):232– 235. PMID:32267343

95. Baumeister RF, Leary MR. The need to belong: desire for interpersonal attachments as a fundamental human motivation. Psychol Bull 1995 May;117(3):497–529. PMID:7777651

96. Gao J, Zheng P, Jia Y, Chen H, Mao Y, Chen S, Wang Y, Fu H, Dai J. Mental health problems and social media exposure during COVID-19 outbreak. PLoS One 2020 Apr 16;15(4):e0231924. PMID:32298385

97. Best P, Manktelow R, Taylor B. Online communication, social media and adolescent wellbeing: A systematic narrative review [Internet].Children and Youth Services Review. 2014. p. 27–36. [doi: 10.1016/j.childyouth.2014.03.001]

98. Berryman C, Ferguson CJ, Negy C. Social Media Use and Mental Health among Young Adults [Internet]. Psychiatric Quarterly. 2018. p. 307–314. [doi:10.1007/s11126-017-9535-6]

99. Pappa S, Ntella V, Giannakas T, Giannakoulis VG, Papoutsi E, Katsaounou P. Prevalence of depression, anxiety, and insomnia among healthcare workers during the COVID-19 pandemic: A systematic review and meta-analysis. Brain Behav Immun 2020 Aug;88:901–907. PMID:32437915

100. Saha K, Torous J, Ernala SK, Rizuto C, Stafford A, De Choudhury M. A computational study of mental health awareness campaigns on social media. Transl Behav Med 2019 Nov 25;9(6):1197–1207. PMID:30834942

101. Lublin N. Notes On Coronavirus: How is America Feeling? Part 7 [Internet]. CrisisTextline. [cited 2020 Jul 25]. Available from: https://www.crisistextline.org/mental-health/notes-on-coronavirus-how-is-america-feeling-part-7/

102. Miltimore J. “A Year”s Worth of Suicide Attempts in Four Weeks’: The Unintended Consequences of COVID-19 Lockdowns [Internet]. Fee.org. [cited 2020 Jul 25]. Available from: fee.org/articles/a-years-worth-of-suicide-attempts-in-four-weeks-the-unintended-consequences-of-covid-19-lockdowns/

103. Seaman J. Suicides in Colorado dropped 40% during first 2 months of coronavirus pandemic but calls to crisis line spiked [Internet]. Denver Post. [cited 2020 Jul 25]. Available from: denverpost.com/2020/05/23/colorado-suicides-dropped-coronavirus-pandemic-calls-to-crisis-line-spiked

104. Jacobson NC, Lekkas D, Price G, Heinz MV, Song M, James O’Malley A, Barr PJ. Flattening the Mental Health Curve: COVID-19 Stay-at-Home Orders Are Associated With Alterations in Mental Health Search Behavior in the United States (Preprint) [Internet]. [doi: 10.2196/preprints.19347]

105. Mackey T, Purushothaman V, Li J, Shah N, Nali M, Bardier C, Liang B, Cai M, Cuomo R. Machine Learning to Detect Self-Reporting of Symptoms, Testing Access, and Recovery Associated With COVID-19 on Twitter: Retrospective Big Data Infoveillance Study. JMIR Public Health Surveill 2020 Jun 8;6(2):e19509. PMID:32490846

106. Stokes DC, Andy A, Guntuku SC, Ungar LH, Merchant RM. Public Priorities and Concerns Regarding COVID-19 in an Online Discussion Forum: Longitudinal Topic Modeling [Internet]. Journal of General Internal Medicine. 2020. p. 2244–2247. [doi: 10.1007/s11606-020-05889-w]

107. Guntuku SC, Sherman G, Stokes DC, Agarwal AK, Seltzer E, Merchant RM, Ungar LH. Tracking Mental Health and Symptom Mentions on Twitter During COVID-19. J Gen Intern Med [Internet] 2020 Jul 7; PMID:32638321

108. Imran M, Castillo C, Diaz F, Vieweg S. Processing Social Media Messages in Mass Emergency [Internet]. Companion of the The Web Conference 2018 on The Web Conference 2018 - WWW’18. 2018. [doi: 10.1145/3184558.3186242]

109. Olteanu A, Castillo C, Diaz F, Kiciman E. Social Data: Biases, Methodological Pitfalls, and Ethical Boundaries [Internet]. Frontiers in Big Data. 2019. [doi: 10.3389/fdata.2019.00013]

110. Watts DJ. Common sense and sociological explanations. AJS 2014 Sep;120(2):313–351. PMID:25811066

111. George Floyd: What happened in the final moments of his life [Internet].BBC. [cited 2020 Jul 25]. Available from: http:bbc.com/news/world-us-canada-52861726

